# Meta-analysis of COVID-19 patients to understand the key predictors of mortality in the non-vaccinated groups in remote settings

**DOI:** 10.1101/2022.01.04.21267659

**Authors:** Neha L. Jain, Karishma Parekh, Rishi Saigal, Amal Alyusuf, Gabrielle Kelly, Alokkumar Jha

## Abstract

Various studies have looked into the impact of the COVID-19 vaccine on large populations. However, very few studies have looked into the remote setting of hospitals where vaccination is challenging due to social structure, myths, and misconceptions. There is a consensus that elevated inflammatory markers such as CRP, ferritin, D-dimer correlate with increased severity of COVID-19 and are associated with worse outcomes. In the present study, through retrospective meta-analysis, we have looked into ∼20 months of SARS-COV2 infected patients with known mortality status and identified predictors of mortality concerning their comorbidities, various clinical parameters, inflammatory markers, superimposed infections, length of hospitalization, length of mechanical ventilation and ICU stay. Studies with larger sample sizes have covered the outcomes through epidemiological, social, and survey-based analysis—however, most studies cover larger cohorts from tertiary medical centers. In the present study, we assessed the outcome of non-vaccinated COVID 19 patients in a remote setting for 20 months from January 1, 2020, to August 30, 2021, at CHI Mercy Health in Roseburg, Oregon. We also included two vaccinated patients from September 2021 to add to the power of our cohort. The study will provide a comprehensive methodology and deep insight into multi-dimensional data in the unvaccinated group, translational biomarkers of mortality, and state-of-art to conduct such studies in various remote hospitals.

## Introduction

The disease caused by severe acute respiratory syndrome coronavirus 2 (SARS-CoV-2), known as coronavirus disease 2019 (COVID-19), has been classified as a pandemic by the World Health Organization (WHO) by February 2020. As of today, over ∼28.7 million cases have been reported worldwide, with more than ∼5.1 million deaths^1^. The effects of the pandemic have been incredibly profound in the unvaccinated group, and a majority of them are destined to comprise the high-risk group of patients^2^. The likelihood of their increased severity is induced by but not limited to advanced age, underlying comorbidities, and compromised immune status. The aforementioned comorbidities can have similar gravitas of disease impact in the vaccinated group; however, the mortality rate is much lower than earlier. One of the proposed mechanisms for increased severity is endothelial dysfunction, most commonly seen in these chronic illnesses and is exacerbated in COVID-19 infection. It is associated with increased complications such as Acute respiratory distress syndrome (ARDS), coagulation imbalance, cytokine storm, and multiorgan failure^3^. It remains elusive if there is a host hyperimmune reaction or host immune dysfunction when infected with SARS-CoV-2, resulting in multi-organ dysfunction. Cytokine storm-related to COVID-19 is associated with poor outcomes. Various cytokine levels such as interleukin-1β, interleukin-6, TNF, and many more are noted to be elevated. It is unclear if the cytokine storm drives COVID-19 infection or is a secondary process.^4^

As discussed earlier, a plethora of studies have indicated a relatively high risk of severe COVID-19 in the unvaccinated group of patients^4^; however, the studies have failed to capture its impact in remote locations and corresponding medical centers with low vaccination rates, which favors the rapid spread of the virus, posing a health risk for all the individuals within the community. Further, most studies in this area have been either behavioral or observational rather than clinical and retrospective to explain the conditions and microenvironment due to remote location challenges. Additionally, previous publications of outcomes for the unvaccinated cohort repeatedly demonstrated relatively high mortality for the subgroup of patients without vaccination.

Given the general paucity of information regarding factors associated with severe COVID-19 in patients without vaccination, relatively high-level information regarding the outcomes of patients in remote settings would be of high interest to the community of remote medical centers in guiding management decisions. In this vein, we have decided to conduct this unique study by performing a descriptive meta-analysis in a remote hospital situated in Roseburg, Oregon. The primary purpose of the current research is to develop a formal statistical model to compare the independent associations of any differences in descriptive and clinical data with fatal COVID-19 outcomes stratified based on the vaccination status. Having such a study conducted in a remote setting hospital will be a beacon of hope for other similar studies and observations in other remote hospitals within the United States and around the world.

To achieve this, we have developed a mathematical model to identify the severity and predictors of mortality in unvaccinated COVID-19 patients trading off clinical variables such mechanical ventilation duration, severe illness requiring admission to a hospital, requirement of ICU level of care, and laboratory values. The primary focus was to build independent associations of outcome/mortality with state of disease, line of therapy received, and recent use of immunosuppressant therapy.

## Background

Most studies on COVID-19 so far have focused on urban centers resulting in a paucity of data on how the current pandemic has affected the rural population and medical centers in the US and across. More diverse studies pertaining to localized information in remote hospitals are needed for COVID-19 infection related to biomarkers, treatments/management, and interventions and their outcomes. To captivate that, we have conducted a study at CHI Mercy Health, a private, not-for-profit 174-bed rural medical center located in Roseburg, Oregon. It has a population of 23,479, of which 22.3% are persons of age 65 and over. About 12.7% of the population live in poverty, with 22.9% on Medicaid and 14.1% on Medicare. The socio-economic conundrum made the COVID-19 impact worse in the area owing to various factors like lack of infrastructure, early treatment unavailability, and failure in convincing local people to get vaccinated. As of October 29th, 2021, the state epidemiological data suggested that only 49% of Douglas county’s population has been vaccinated. Low rate of vaccination added a lot more to the emotional, social, and personalized health burden on the medical fraternity by in-patient hospitalization flow due to low vaccination rate, which led the resources under severe strain^5^. Various studies also looked into the retrospective data to understand the clinical and social biomarkers to educate the community regarding better planning and management of the disease. Various published studies have helped build models based on identifying risk factors to predict severity and mortality and assist the hospitals in better preparing for the COVID-19 surge. Thus, the proposed study intended to review the medical records of deceased COVID-19 patients in our medical center to understand the disease progression during their respective hospital courses. The analysis of various variables, including demographics, medical comorbidities, clinical data, medical treatments, and their effects on each other, is expected to elucidate the COVID-19 disease, predict severity and understand the mortality trend. It is critical to conduct such a study especially in the remote setting hospital, to obtain the pattern despite complex patient heterogeneity. For example, as per **(Supplementary Table S1)**, patient #24 is an unvaccinated female in 70s with a history of hypertension, was found to be hypoxemic at 89%, requiring 15 lpm of supplemental O2. She had severe COVID-19 with imaging findings consistent with bilateral infiltrates, laboratory values confirming lymphopenia, elevated LDH, and D-dimer requiring prolonged ICU course and mechanical ventilation. She was treated with remdesivir, steroids, paralytics, and antibiotics for bacterial pneumonia. Her course was complicated with pulmonary embolism, ARDS and renal dysfunction. She underwent tracheostomy but eventually was transitioned to comfort care. Another patient #35 is a female in mid 60s with a history of a liver transplant, obesity, hypertension, diabetes mellitus, chronic kidney disease who presented with SpO2 of 82% on room air. She had a prolonged length of stay complicated with mechanical ventilation, ARDS, pulmonary embolism, shock, and fungal pneumonia, eventually transitioned to comfort care. She was appropriately treated with steroids, remdesivir, anticoagulation, antibiotics, proning, paralytics, and antifungals. Her imaging showed moderate bilateral infiltrates, and labs revealed elevated LDH and lymphopenia. The above examples of unvaccinated and vaccinated patients (one each) show the complexity in the patient presented in terms of their comorbidities and age. It is essential to associate the vaccination status with other parameters to identify the predictors of mortality that can explain the vaccination as a multivariate model. For example, closely looking into our cohort, including the above two patients, obesity reveals as one of the critical comorbidities in COVID 19 patients. This has also been reported in many other studies, including a CDC report which suggests that approximately 40% of US adults are obese. Similarly, some demographic data have suggested that the risk of severity, intensive care admission, and mortality increases with higher BMI^9^. Seeing this pattern, we identified inflammation markers (detailed discussion in the results section) as critical predictors of mortality more precisely in the unvaccinated group. One such critical marker is D-dimer, one of the fragments produced during the degradation of blood clots and is known to be elevated in patients with moderate to severe COVID-19. The elevation of D-dimer values correlates with disease severity and is attributed to be a prognostic marker for inpatient mortality^6^. Similarly, another clinical marker that can be critical predicting poor outcomes is lymphopenia. It is associated with severe COVID-19 pneumonia and higher mortality. The mechanism of lymphopenia in COVID-19 pneumonia remains obscured. There are hypotheses such as lymphocyte sequestration in lungs, gastrointestinal tract, and lymphoid tissues versus ACE receptors on lymphocytes being a direct target of SARS-CoV-2 versus Cytokine storm further reducing the lymphocyte count ^7–10^.

Despite the plethora of research another critical aspect that remains warranted and requires more accumulation of clinical findings is the need for supplemental oxygenation in COVID-19 patients; higher the need for supplemental oxygen, the higher the risk for severe COVID-19 and death. This will guide the initial placement of the patient appropriately among intensive care unit, step down unit/progressive care unit, medical floors and monitor them closely for decompensation^10^.

Looking through the clinical and observational parameters in deceased unvaccinated and vaccinated COVID-19 patients, it appears that point several common factors point toward the worst outcomes. These subgroups of patients presented with hypoxemia on arrival had elevated inflammatory markers, lymphopenia, bilateral lung involvement, required mechanical ventilation, and had a prolonged hospital stay. These patients often developed complications such as ARDS, multi-organ dysfunction, superimposed infections, pulmonary embolism, pneumothorax, and pneumomediastinum during their hospital stay, eventually leading to the same outcome of death.

One concluding factor that can have both logistic and clinical impact in remote settings, hospitals, and low-income areas is the length of stay (LOS). Various serological studies (including our cohort) suggested that severe COVID-19 patients develop acute respiratory distress and multi-system organ failure and are associated with poor prognosis and higher mortality. They have associated the severity with the length of ICU stay and hospitalization ^11–13^. For instance, Li et al ^14^ suggested that based on severity defined through inflammatory markers, the length of hospitalization stay was ten days (IQR median) in all the patients, 12.5 days in severe disease (including critically ill), nine days in mild to moderate disease. This is also reciprocated in our cohort, where the length of hospitalization is 11.0 days in the vaccinated group and 13.5 days in the unvaccinated group. Thus, the correlation of LOS with other critical markers will barter as a prognostic evaluation model for critically ill patients in remote settings.

The above mentioned explains the prognostic and clinical presentation of these biomarkers. To summarize, our retrospective analysis aims to provide a bird’ eye view of uniqueness and possible disparity, if any, in a rural community-based hospital. The outcomes may help the medical community in rural areas assess, stratify, and closely observe high-risk patients and prevent untimely death.

## Materials and Methods

### Statistical Analysis Plan

The primary model will be a logistic regression with the outcome variable being the composite endpoint of severe COVID-19. Multiple imputations will be performed on missing variables. For the secondary analysis of mortality, logistic regression based on 30-day mortality will be served. The goodness of fit of the models will be assessed by Harrell’s C-statistic with a 95% confidence interval determined by the method of DeLong. Variance inflation factors will be computed for every predictor in the model. We plan to conduct sensitivity analyses restricting the primary research to patients with primary composite endpoint, omitting one component of the severe COVID-19 composite endpoint (severe illness requiring hospitalization). The exact process for the preceding model will be conducted for the sensitivity analyses. All analyses will be performed using R v3.6.3, packages “rms”, “Hmisc”, “rpart”, and “train.”

### Data Collection

All of the variables requested for this analysis are either directly coded by the medical records group at Mercy Medical Center, Roseburg, or further investigators derived variables utilized in the course of analysis. Thus, obtaining the information necessary to complete this proposal should be feasible; however, due to hospital data guidelines, the data will only be available upon request after signing the Data Transfer Agreement (DTA).

Regarding the modeling exercise, the size of the cohort and rates of outcomes allowed us to complete the analysis as proposed. As of August 31, 2021, there were 38 patients with confirmed vaccination status, 5 of whom met the composite endpoint of severe COVID-19 in the vaccinated group. The model for our primary analysis involves a total of 36 degrees of freedom: age (1), sex (1), lab data performance status category (2, ordinal variable with 3 levels: 0, 1, 2 or greater), smoking status (1, binary variable, current/former or never), state of disease (2, a categorical variable with 3 levels: date of the first admission,.), line of therapy received (2, categorical variable with 3 categories: untreated, first line, second line), phase of therapy (2, categorical variable with three categories: induction, consolidation, maintenance), and recent receipt of cytotoxic therapy (1, binary variable). Therefore, the number of events is sufficient to power this analysis, assuming 26 events per degree of freedom. Additional patient characteristics are demonstrated in **Table 1** and **Table 2**.

**Table 1:** The clinical characteristics of the COVID-19 patients during the 20-month period, based on the mean (IQR)

**Table 2:** The clinical characteristics of the COVID-19 patients were stratified by vaccination status (vaccinated, unvaccinated, and unknown) over the 20 months, based on the mean (IQR). The Kruskal-Wallis rank-sum test determined the significance of distribution, Fisher’s exact test.

### Sensitivity analyses

Analyses were performed in R version 4.0.3 (R Foundation for Statistical Computing, Vienna, Austria), including the Hmisc, rms, ordinalNet, UpSetR, mice, extension packages.

### Missing data Imputation

As shown in **(Supplementary Table S1)**, we had missing values for several clinical variables such as D-Dimer (0-0.52 ng/l), Lymphocyte count (%), etc. Given the small sample size of the study, we have an imputation algorithm to predict the missing values; we have used the R package “mice” and “VIM” to visualize and predict the missing values. We have used the k-NN algorithm with k=6 (Square root of the total number of patients) to determine accuracy. We have identified only two patients with different predicted values of D-dimer and Lymphocyte count. We have used values predicted through “mice” as final values for the classification to identify the predictors of mortality. We have used m=5, maxit=50, meth=‘pmm’, seed=500 to calculate the missing value in mice function. As shown in (**Figure S2(a))**, we have missing values in several variables, and (**Figure S2(b))** shows post imputation; we didn’t have any variables. The list of complete data is listed in **(Supplementary Table S2)**.

### Power calculation

The sample size of 38 patients (32 Vaccinated, 5 Unvaccinated, 1 Unknown) accrued as of August 2021 yields 80% power at an alpha of 0.05 to detect a hazard ratio of 0.01 for severe COVID-19, assuming two unequally matched groups.

### Classification and Predictors of mortality

Vaccinated, unvaccinated, and undetermined groups were used to build a model and identify the variables critical to the non-vaccinated group. We used “caTools’” and “randomForest” R package to classify and preprocess the data. Our model was trained using 75 % of the patients and tested using 25%. To split the data, we used the caTools split function. We have used mtry=6 and No of tress= 501 to train the model. Gini Score has been used to determine the 15 top variables.

### Logistic Regression

Using the R packages “epitools” and “gtsummary,” **Table 1, Table 2**, and **Table S1** are generated to show patient demographics and a logistic regression equation to evaluate the variables contributing to mortality assess unvaccinated patients.

### Charlson Comorbidity Index (CCI)

We calculated the Charlson Comorbidity Index (CCI) with the following assumptions, firstly for age < 50 we assigned score of 0, age >= 50 (1), age < 60 ∼ 1, age < 70 ∼ 2, age < 80 ∼ 3, age >= 80 ∼ 4. Similarly, for autoimmune, obesity, CHF, HTN, ASCVD, COPD, asthma, ILD, OSA, the availability of events was scored “1” else “0”. For CKD, past cancer, transplant availability of event was score “2” else “0”. Further, for Active cancer availability of event was scored “6” else “0”, followed by DM where availability of event was scored “3” else “0”. We used the r package “comorbidity” to calculate CCI and “ggplot2” to plot CCI.

## Results

In the context of remote hospitals, patients and potential outcomes were distributed in a manner that led to some of the most critical outcomes for COVID 19. The patient characteristics are outlined in **(Figure 2). Table 2** depicts the distribution of patients in our medical center. **(Figure 2)** illustrates how we categorized 71 collected clinical observations and potential biomarkers across eight broad categories. As indicated by **(Figure 2)**, one of the strengths of our data is the prevalence of heterogeneity, such as marital status*, BMI, and smoking status*. Furthermore, as stated earlier, most of our patients are not vaccinated. Numerous studies have suggested that lifestyle and social factors (marital status, smoking)* impact outcomes ^15 16^. Similarly, smoking and passive smoking during the lockdown also negatively impacted children, as well as adults and family members^17^. We had 15 former and two current smokers in our cohort and 23 patients with higher BMIs (>30). In COVID-19 patients^18^ both factors have been associated with the worst outcome. Additionally, chronic diseases such as cancer, autoimmune diseases, cardiovascular disorders, asthma, and hypertension contribute towards the worst outcome in COVID 19 infected patients who were unvaccinated^19^. Also, the cohort is sufficiently extensive to explain the associated complications that may be causing the worst outcome in COVID 19 unvaccinated patients. According to **Figure 2**, we have collected laboratory findings such as CRP, D-Dimer, Lymphocyte, CT-based severity, SpO2 on arrival, treatments, and complications. The selection will allow us to explain the role of biomarkers converges to predictors of mortality and aid the treating physician in opting for further treatment. In light of the factors presented in **(Figure 2)**, the following are significant conclusions we derived from the current study.

**Figure 1:**
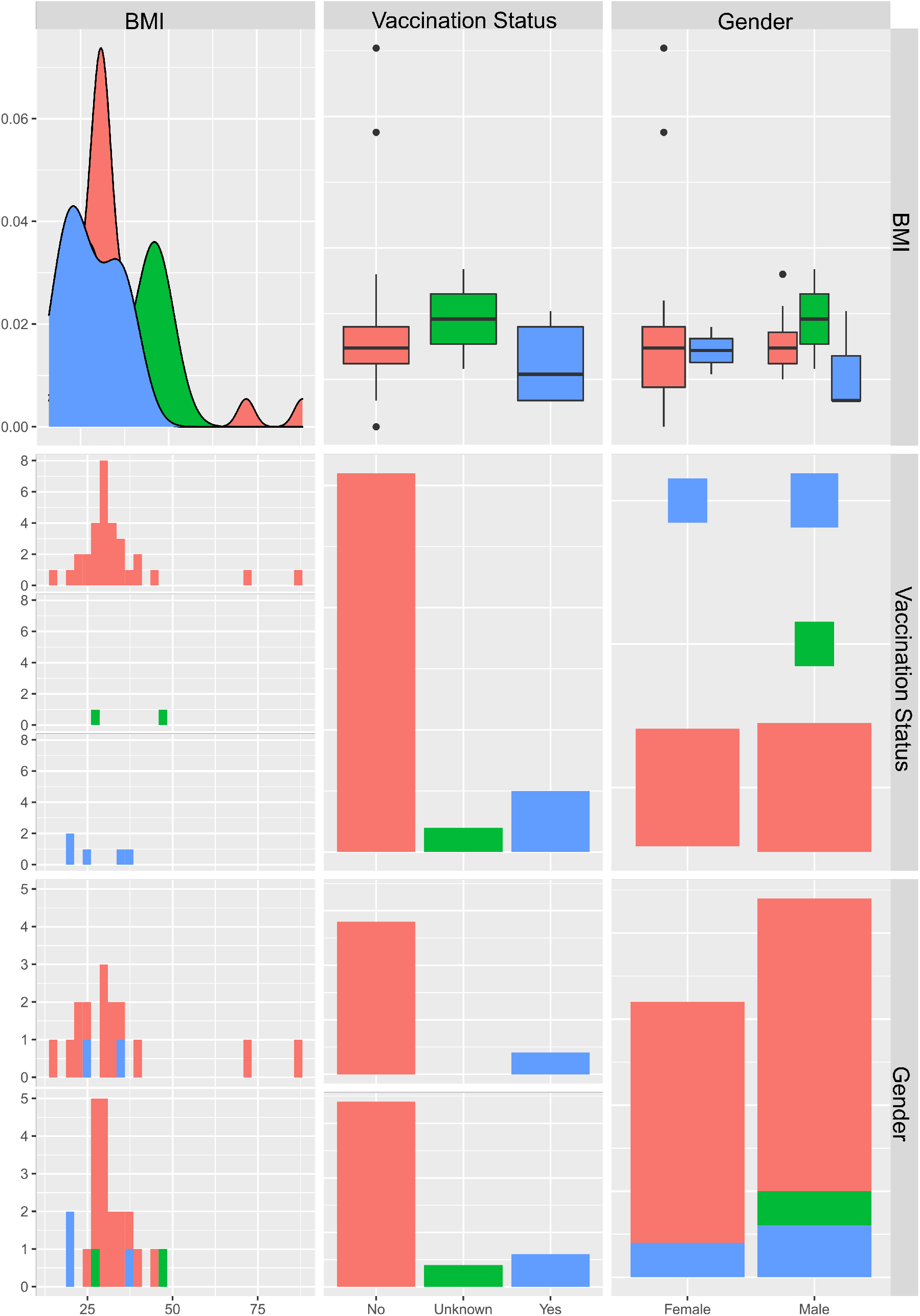
The overall distribution of patients according to their vaccination status. Based on age distribution and BMI, Pearson’s correlation was calculated.

**Figure 2:**
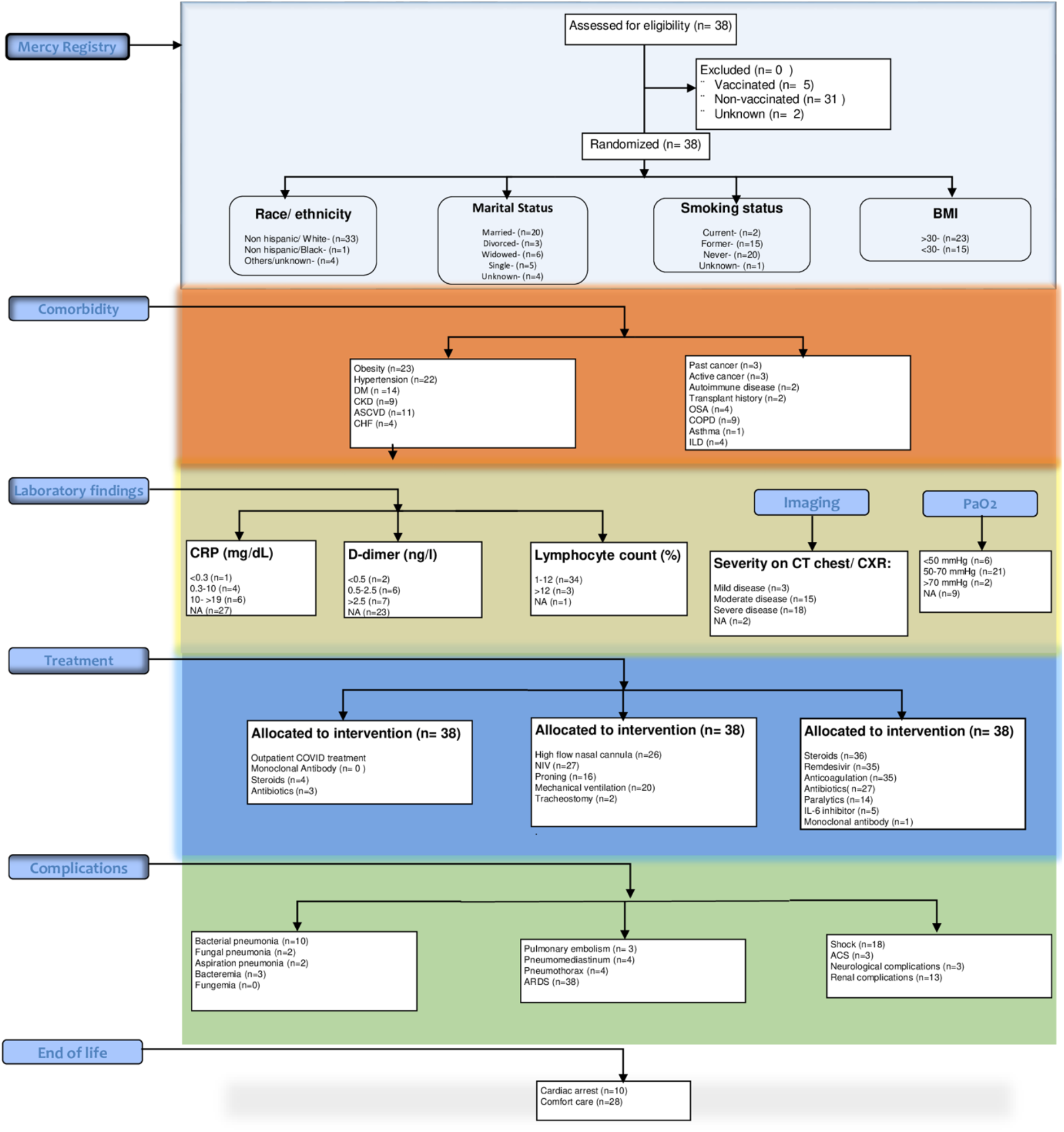
The flow chart indicates all the clinical variables collected and their distribution based on demography, treatment, complications, laboratory findings, and end of life distribution.

* Smoking status, Marital status, Age, Race, Ethnicity, Gender, Weight, Height is subject to IRB approval and available for peer review only. The * sign only denoted at the first occurrence of variable, however, the data availability conditions remain the same for earlier mentioned variables.

### Patient Distribution

Our cohort consisted of 38 records. There were no exclusion criteria applied, resulting in 38 records being included in the primary analysis **(Supplement Table 1)**. The median age of the included cohort was 75.5 years (interquartile range: 44 to 95 years) *, and the majority, 57.89% (n=22), were male, and 42.1 % (n=16) were female*. Non-Hispanic whites accounted for half the cohort, while non-Hispanic blacks* accounted for 2.63% (n=1). 81.57% (n=31) of the patients were without any vaccinations. These are similar proportions to the patients without vaccination, 10.52% (n=4) were treated within first three months, and 89.47% (n=34) were not treated within first three months of data collection. Over 47.36% (n=18) of the cohort had severe bilateral lung involvement based on imaging, and over 52.63% (n=20) required mechanical ventilation. 92.1% (n=35) received remdesivir; 94.73% (n=37) received steroids and 13.15% (n=5) received tocilizumab. A total of 81.57% (n=31) of the patients were unvaccinated. **Tables 1** and **2** provide additional information regarding the patient.

### Patient outcomes

The data collected over the span of two weeks from a remote hospital setting led to critical observations. We calculated interquartile range (IQR) median values for the variables. Thus, all discussion further will be IQR median. As demonstrated in **(Table 1)** in 38 patients, the median age is 76. It is essential to note that our population of patients is distributed as follows: Unvaccinated: 31 (82%), Unknown and Vaccinated are 2 (5.3%) and 5 (13%), respectively. We further looked into some of the critical COVID 19 associated variables in the non-vaccinated group to compare all-cause mortality among severe COVID 19 patients treated at Mercy Medical Center, Roseburg, Oregon. There have been various criteria to define the severity of illness; however, as discussed in recently ^20–23^ published articles following clinical parameters have higher predictive value in hospitalized and unvaccinated COVID 19 patients. a) Respiratory distress, Respiratory rate > 30/min b) Mean O2 saturation ≤93% in the resting state c) partial pressure of oxygen in arterial blood (PaO2)/oxygen concentration (FiO2) ≤300 mmHg d) lung involvement on imaging >50% within 24–48 h e) The critical value of initial D-Dimer, peak D-Dimer, initial NLR (neutrophil to lymphocyte ratio) and peak NLR in prognosticating of intubation was 0.73 mg/L, 12.75 mg/L,7.28 and 27.55 f) lymphocytopenia g) duration of admission post-infection.

#### Lymphocyte count and D-Dimer in conjunction with other clinical characteristics may influence the outcome of unvaccinated COVID 19 patients in remote areas

Earlier, we talked about how the more extensive cohort study analysis can facilitate the potential biomarkers for the worst outcome in COVID 19. **Figure 1** illustrates how BMI, vaccination status, and gender are distributed and suggests that age, gender, and body mass index (BMI) may impact the immune response to vaccines^24^. Furthermore, the epidemiological studies within the state of Oregon demonstrate that 30% of the adults in Douglas County are obese, which is significantly higher than the state average of 27%, despite the county having an average age of 38.7 years^25^. According to the County Vaccine Tracker, as of 19th November 2021, the vaccination rate stands at 55.3%, and the risk level is extremely high. To account for this, we examined our cohort’s age, gender, BMI, and vaccination status **(Figure 1)**. As shown in **(Figure 1)**, BMI is relatively higher in unvaccinated male patients. For the sake of simplicity in the study, we avoided conclusions for the unknown vaccinated group due to their singleton number. The male gender was also associated with higher mortality in earlier studies^26^. Therefore, we can correlate our indicators and predictors of mortality with larger-scale studies despite the small cohort size. Once we established the social parameters in the earlier section and their role in the outcome, we also looked into all the clinical variables to identify the predictors of mortality to differentiate the vaccinated and unvaccinated patient groups. As discussed and explained in the methods, we used binary classification (vaccinated, unvaccinated) using Random Forest (RF) machine learning method to classify the groups. Further, we used *Gini Score* to rank the clinical variables. The ranked list (Top 15) of clinical variables that differentiate groups in all patients with the worst outcome is depicted in **Figure 3(a)**.

**Figure 3:**
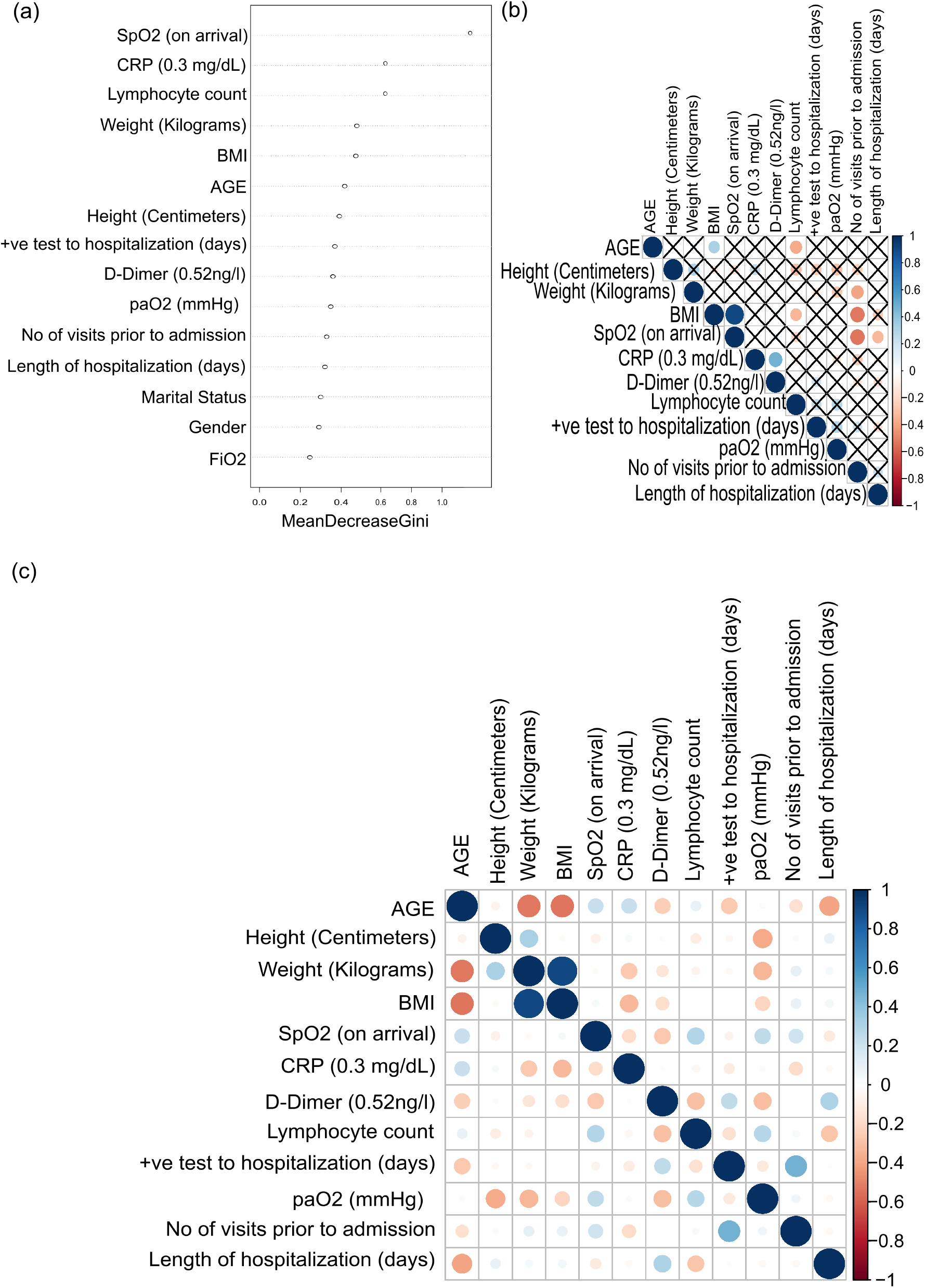
(a) Shows top 15 clinical variables based on random forest classification Gini score. (b) indicated the correlation between the top 15 clinical variables. The cross shows the non-significance based on p-value < 0.01. (c) defines the correlation among the top 15 clinical variables without indicating p-value significance. It shows that the correlation and p-value can be omitted due to the non-heterogeneous small sample size.

Further, due to our small sample size, we also looked into the conservative approach of identifying the correlation between different variables and extracting the significant variables that may have contributed to mortality **Figure 3(c and b)**. As demonstrated in **Figure 3 (a**), SpO2 (on arrival), CRP level, lymphocyte count, BMI and associated markers (age, weight), length of hospitalization, D-dimer, and paO2 (mm/hg) are expected to be predictors of mortality in unvaccinated patients. For a closer look at these variables, the correlation plot illustrates the correlation between them. As shown in **Figure 3 (b)**, age, BMI, and lymphocyte count have a negative correlation of outcome, and it has been established that lymphocyte count, B cells, NK cells, cytotoxic and helper T cells are reduced in severe COVID 19 patients^27^.

The severity signifies the inverse relationship and elucidates the severity of SARS-COV2 in our cohort. Further, weight (kilograms), BMI, and CRP levels also negatively correlate with the outcome for the number of visits prior to final admission. As shown in **Figure 3 (a)**, these markers are elucidated as critical predictors of mortality, and a higher number of visits prior to admission can correlate with delay in treatment and response. Studies have shown that weight (kilograms), BMI, and CRP levels are predictors of mortality in severe COVID 19 cases. It has been documented that severe COVID 19 is associated with higher inflammatory mediators due to cytokine storms. These markers may allow early identification or even prediction of disease progression and define admission priority^28.^ CRP is also known as an active regulator for innate immunity and predictor for mechanical ventilation. According to the positive correlation between CRP and D-dimer values, both of these values are likely to be elevated during the early stages of COVID 19 infection due to systemic inflammation^29,30^. Both these values have a proportional relationship with the severity of the disease. As discussed earlier, similar to CRP, elevated D-dimer at the time of admission could be an indicator of severe COVID 19 infection requiring ICU admission^31^ as shown in **Figure S3 (b)**. All our patients had elevated D-dimer during treatment compared to the vaccinated group (actual and predicted) displayed in **Figure 2** and **Table 1**. Further, D-dimer levels on hospital admission have been identified as being associated with increased mortality, and a positive correlation of CRP and D-dimer is also associated with cardiac arrest and coronary heart disease. As we can see in **Figure 2, Table 1, and Table 2**, we had a higher number of unvaccinated patients with a high prevalence of cardiac arrest.

The elevation of CRP and D-dimer also increases the probability of complications such as pulmonary embolism, acute respiratory distress syndrome (ARDS), and superimposed bacterial pneumonia^32^. **(Figure 2)** and **Table 2**, indicates that many unvaccinated patients from our small cohort had these complications. Hence, it is established that endothelial damage caused by inflammation leads to increased severity of COVID 19 in our patient cohort. The systemic understanding and function are discussed below in the discussion section. However, even with the remote setting, the systemic track of CRP level, D-dimer, and lymphocyte count can allow the physician to plan for early hospitalization and mechanical ventilation. The overall association of correlations among a few clinical variables that may not be statistically significant due to the small sample size is also depicted in **Figure 3 (C)**. As such, **in Figure 3(B)**, SpO2 (on arrival) has a strong negative correlation of outcome with the number of visits prior to final admission and length of hospitalization (days). Precisely, oxygen saturation (SpO2) values of less than 90% on admission have been identified as the predictor of mortality in COVID 19 patients. Another strong correlation was seen between PaO2/FiO2 & SpO2/FiO2. Even though not evident based on the correlation plot as shown in **Figure 3(a)**, both are critical variables to differentiate vaccinated and unvaccinated groups in our machine learning model. The significance proves that PaO2/FiO2 is a strong predictor of mortality, as shown in **Figure 3(c)**. Even though values are not statically significant, PaO2 and SpO2 positively correlate with the number of visits prior to final admission. We did not see any solid indicators for the length of hospitalization, SpO2, and PaO2 despite being critical predictors of mortality in COVID 19 patients^33^. However, these two factors can be one of the mediatory factors in the putative connection between the length of hospitalization, number of visits prior to final admission. Hence, we elucidate that despite limited resources, many such studies can be conducted in remote centers to understand the location-agnostic prognostic tool of COVID 19 management and treatment, such as hospitalization based on vaccination status and other treatment options.

#### Age and other metabolic features contributed highly to mortality along with inflammatory markers

We calculated the Charlson Comorbidity Index (CCI)^34^ to identify the pattern of mortality and contributing factors that may be causing the higher levels of inflammation. According to **(Figure S5)**, the distribution of CCI across all comorbidities in our cohort clearly illustrates that most patients fall into the high-risk group with higher age, BMI, and obesity. Additionally, hypertension and diabetes are prevalent, indicates that metabolic pathways played a significant role in mortality. Numerous studies have suggested that patients with COVID 19 have higher levels of cytokine storms due to cardiometabolic disease^35 36^. The Centers for Disease Control and Prevention (CDC) contends that Douglas county is at greater risk for COVID 19 due to its average age, diabetes, and high body mass index (BMI). Additionally, we examined individual CCI for each patient, as shown in **(Figure S6)**. Keeping the CCI score zero as a baseline, it is evident that any score in the range of three is prognostically indicative of death or poor outcome in COVID 19 patients. An increase of one point above this will result in an increase of 16% in mortality. **(Figure S6)** depicts that most of our patients had CCI > 3, which indicates that most of these patients were considered high risk. M2 and M16 with higher CCI were unvaccinated, while M36 with the lowest CCI was vaccinated. Even the lowest CCI patient was >3, meaning that this patient was in a higher-risk group with obvious signs on imaging and hypoxemia on presentation (SpO2 of 80% on room air).

## Discussion

COVID 19 poses unique risks to patients without vaccination. This study examined the characteristics and clinical outcomes of a small cohort of COVID 19 unvaccinated and vaccinated patients in a remote hospital in Roseburg, OR, USA. Unvaccinated Non-Hispanic/ White-male patients represented the majority of all COVID 19 positive patients who could not survive post-treatment or hospitalization. Patients had a higher prevalence of obesity, hypertension, diabetes at baseline. A higher percentage of unvaccinated patients presented with elevated inflammatory markers levels than unknown and vaccinated patients. Most of the patients who received critical care or mechanical ventilation were unvaccinated and obese. The white race, higher median age, a higher score on the Charlson Comorbidity Index, public insurance (Medicare or Medicaid), resident in a low-income area, remote location of the hospital, limited healthcare resources, higher inflammatory markers, and obesity were associated with increased odds of hospital admission **(Table 2)**. Whites (Male) were overrepresented among all patients who died in the hospital (78.6%). The current trends in Douglas county^1^ being catalogued as a high-risk zone insinuate that lower vaccination rates and comorbidities prevalent in the area are expected to be the prognostic predictors of mortality. The remote settings also have their challenges (see next section) and other social influences indicated by CDC, such as a higher rate of vaccine hesitancy and dependence on public insurance. This outcome is in line with earlier published studies with similar results^37,38^. Hypoxemic respiratory failure, acute respiratory distress syndrome, superimposed pneumonia, renal dysfunction, and shock were the most commonly identified complications during hospitalization. Relevant laboratory abnormalities included lymphopenia, elevated levels of D-dimer, and other markers of inflammation.

As described in the results **Figure 3(b)**, CRP, D-dimer, lymphocyte count, BMI, and PaO2/FiO2 ratio are the key predictors of hospitalization and mortality. In addition, our results revealed that respiratory deterioration-associated biomarkers (PaO2/FiO2 and SpO2 on arrival) have a higher predictive capability of mortality.

We have conducted a single-center and remotely located hospital’s retrospective cohort analysis of deceased COVID 19 patients. We started teasing out the data from an observational perspective, as displayed in **(Table 1)**. We have used median (IQR) to generate all the summaries and confidence intervals during analysis^39 40^. A higher wait time for care among low-income patients may also result from the severity of their clinical presentation with multiple ER/urgent care visits before hospitalization. Several factors may contribute towards the occurrence of COVID 19 in our study population, including remoteness, disparities in vaccination coverage, and other factors, suggesting that death in our population are likely multifactorial. In addition, the lower-income level, hesitation regarding vaccinations, and lack of access to upgraded medical facilities may have adversely affected their admission time.

According to the Oregon State Public Health Department^2^, Douglas county hospitals are still stretched thin in providing care to COVID 19 patients; most are unvaccinated. In addition, the hospital continuously faced staff and bed shortages causing the grinning effect. The report also indicated that obesity, diabetes, and hypertension were more prevalent in low-income and lower education counties, irrespective of race.

Our cohort found that elevated levels of CRP and D-dimer correlated with disease severity. It is established that COVID 19 is associated with a high incidence of thrombotic complications due to the unique interplay between the SARS-COV2 virus and endothelial cells. The D-dimer molecule is a product of the degradation of the fibrin protein after lysis by the thrombin, plasmin, and factor XIIIa enzymes. This method can also be used in clinical evaluation to identify ongoing coagulation. A higher CRP level induces endothelial dysfunction as well.

This study confirms previously described clinical presentations, laboratory findings, and outcomes of COVID 19–related hospital admissions. The observed differences in the clinical presentation may also reflect differences in underlying chronic conditions on hospital presentation. To explain the relationship of the clinical presentation, we had a non-parametric correlation analysis (**Figure S3(c)**), the positive correlation between the number of days on mechanical ventilation, the number of days in ICU, and the length of hospitalization are unique to our cohort. Symptoms onset prior to arrival and positive tests to hospitalization also have a positive correlation. Our findings suggest that more studies are warranted to assess the response of inflammatory markers in response to this novel coronavirus with respect to low-income status, distance from medical centers, and vaccination status. These and other unrevealed factors may influence the difference in the rate of hospitalization, the timing of patients’ admission, the rate of increasing hospital’s capacity, and the overall outcome.

This study mirrored the findings of similarly conducted studies that showed associations between the risk of in-hospital death, demographic factors (age, BMI), clinical factors (D-dimer, CRP) as well as other biomarkers (as per **Figure 1(a)**). For example, in a meta-analysis of Deng et al., ^41^ Lodigiani et al.^30^ suggested that the lowest to the highest increase in D-dimer was more strongly associated with death than patients with lower D-dimer at baseline. In addition, a positive correlation was found between CRP levels and the need for supplemental oxygenation and invasive ventilation. This pattern was also evident in our small cohort. It has been suggested in a number of studies, including Hu et al. ^6^, and Srikant et al. ^42^ that CRP, D-dimer, and lymphocyte count can be used as prognostic markers of disease severity for COVID 19 patients.

Despite the limited sample size, difficulties in obtaining data, remote infrastructure, and missing data, our statistically valid methods permitted us to conclude that D-Dimer and CRP are prognostic biomarkers in any setting and correlate with the need for mechanical ventilation is excellent. Moreover, this information has also concluded that inflammation-induced cytokine storm is associated with coagulopathy in COVID 19 infected patients. Early rise in D-dimer suggests that coagulopathy acts as a prodrome of cytokine Nonetheless, it was challenging to see any direct effects of treatment, we followed the standard treatment protocols as described in a variety of exciting articles^43^, and our observations can be seen in **(Figure S4)**. In terms of treatment, the mean hospital stay was 11.4 (M) and 9.8 (F) days, respectively. Most commonly prescribed medications were dexamethasone (n=37), remdesivir (n=35), enoxaparin/UFH (n=35) and tocilizumab (n=5). As indicated in **(Figure S4)**, 20 patients required mechanical ventilation, 27 required non-invasive ventilation, and 26 required high-flow oxygen through nasal cannulas. According to our data in **Table 1** and **Table 2**, most of our patients had complex comorbidities **(Figure S5)**, with medications and interventions showing minimal impacts. Many studies have reported that low-risk patients with mild COVID 19 infection, stratified based on D-Dimer and CRP levels, can be managed symptomatically without any specific drug intervention. Consequently, these markers can also monitor progression in extremely critical care patients. Thus, our biomarkers can also be utilized by the community to a) determine an individual’s i) develop a treatment plan, (ii) determine the need for hospitalization, and (iii) eligibility for mechanical ventilation.

Although this study was able to identify critical biomarkers for severe COVID 19 patients, it also has some limitations. This being a retrospective analysis, the findings are primarily used in generating hypotheses, not necessarily in clinical practice. Furthermore, as a registry analysis, there is an inherent bias in the patient selection determined by registry entrant decided based on pre-defined criteria such as outcome in our case. Consequently, the study population likely has a higher COVID 19 severity than the general population of cancer patients with COVID 19. Additionally, as discussed above, this study included a more significant proportion of unvaccinated patients than other analyses, and this may have led to improved generalizability reflecting the impact of COVID 19 in remote settings. However, to maximize sample size, a small number of vaccinated patients were added to the cohort, limiting the generalizability of the findings. Additionally, there are significant differences between the timing of COVID 19 diagnosis in this study and the epidemiology of the pandemic in the general population, with an overrepresentation of diagnoses in the early part of the pandemic period. In all likelihood, this study does not include patients with delta-variant since it includes data up until the third trimester of 2021. In light of the ordinal outcome of COVID 19 severity, including metrics such as hospitalization and ICU admission, patients without vaccination may have an advantage in being admitted to higher levels of care instead of their intrinsic clinical presentation.

It was observed in our study cohort, patients with severe hypoxemic respiratory failure reported increased length of stay prior to the need for mechanical ventilation. While this may not explain all the parameters linked to increased COVID 19 severity, it may contribute to some findings. Furthermore, our study is the largest to date for unvaccinated patients in remote settings and uses machine learning to demonstrate that inflammation affects COVID 19 severity, the sample size was insufficient to determine the independent effects of specific treatments. Future efforts should be aimed at performing such analyses. Unfortunately, some laboratory investigations were not conducted on all patients. The role of these factors in the clinical presentation of the study population may not have been sufficiently considered.

Based on the results of this analysis, it is concluded that patients without vaccination in remote settings have a relatively increased risk of developing severe COVID 19. This risk is further increased in conjunction with other clinical factors. It is essential to consider the individual patient scenario in the context of community infectivity, hospital resources, patient’s baseline characteristics, and disease response with prognosis. The health care needs of this population remain high, and they require improved primary prevention with therapeutic strategies.

Notwithstanding these limitations, this study provides comparative epidemiological characteristics of unvaccinated patients in remote setting hospitals who are underrepresented in the COVID 19 medical literature to date. The study is also intended to shed light on differences in clinical presentation in such remote hospital settings.

### Limitations in remote setting hospital for health care delivery

In the wake of COVID 19, healthcare workers felt the brunt of it. Often this impact is multidimensional, encompassing factors like work management, mental health, and social needs. Another factor contributing to the prevalence of these issues is the disparity between medical centers in urban and rural areas. For example, the pandemic had twice the mortality rate in remote centers than in urban centers^44^. In addition, a large number of rural centers were dealing with shortages of physicians and limited access to vital lifesaving equipment, including non-invasive ventilators, ventilators, dialysis machines, and medications that are required in large quantities to treat moderate to severe COVID 19 infections.

One more Gordian knot was the scarcity of nurses, respiratory therapists, and medical residents necessary to ensure adequate coverage within the already overwhelmed intensive care units (ICUs) and uninterrupted overflow into the COVID units. Through this uniquely positioned scenario, some of the most pressing issues during the pandemic were highlighted, and respondents were asked to display resilience during overworking conditions worsening their physical and mental turmoil. In part due to hospitals having an uneven distribution of resources, transportation of patients needing higher levels of care, such as the trial of extracorporeal membrane oxygenation (ECMO), was further hampered and prolonged. Moreover, tertiary centers participated in the COVID 19 experimental treatments in a rapidly evolving protocol environment. As a result, several new medications were made available, and FDA compassionate release was granted for their patients. On the other hand, the remote hospitals had limited access to resources and were more vulnerable. Among such scenarios is outpatient monoclonal antibody treatment availability in rural settings with limited infrastructure to potentially reduce the severity of the COVID 19 infection set up late in the pandemic.

Even with these obstacles, the hospital administration endeavors to provide up-to-date health care for their community members. In addition, the advent of telemedicine has enabled urban and rural areas to stay closely connected during this pandemic period. However, the fact remains that many possibilities remain unexplored and yet to be implemented to continue catering quality medical care throughout the country.

### Imputation Impact

We imputed clinical variables with a K-NN algorithm to fix the missing data issue; although theoretically, it was more sensitive and specific, the actual values could be different based on clinical representation, treatment, and other prognostic variables.

## Data Availability

All data produced in the present study are available upon reasonable request to the authors. All the patient number are randomly allocated and anonymized to follow PHI guidelines.

## Supplementary File Details

**Figure S1:**
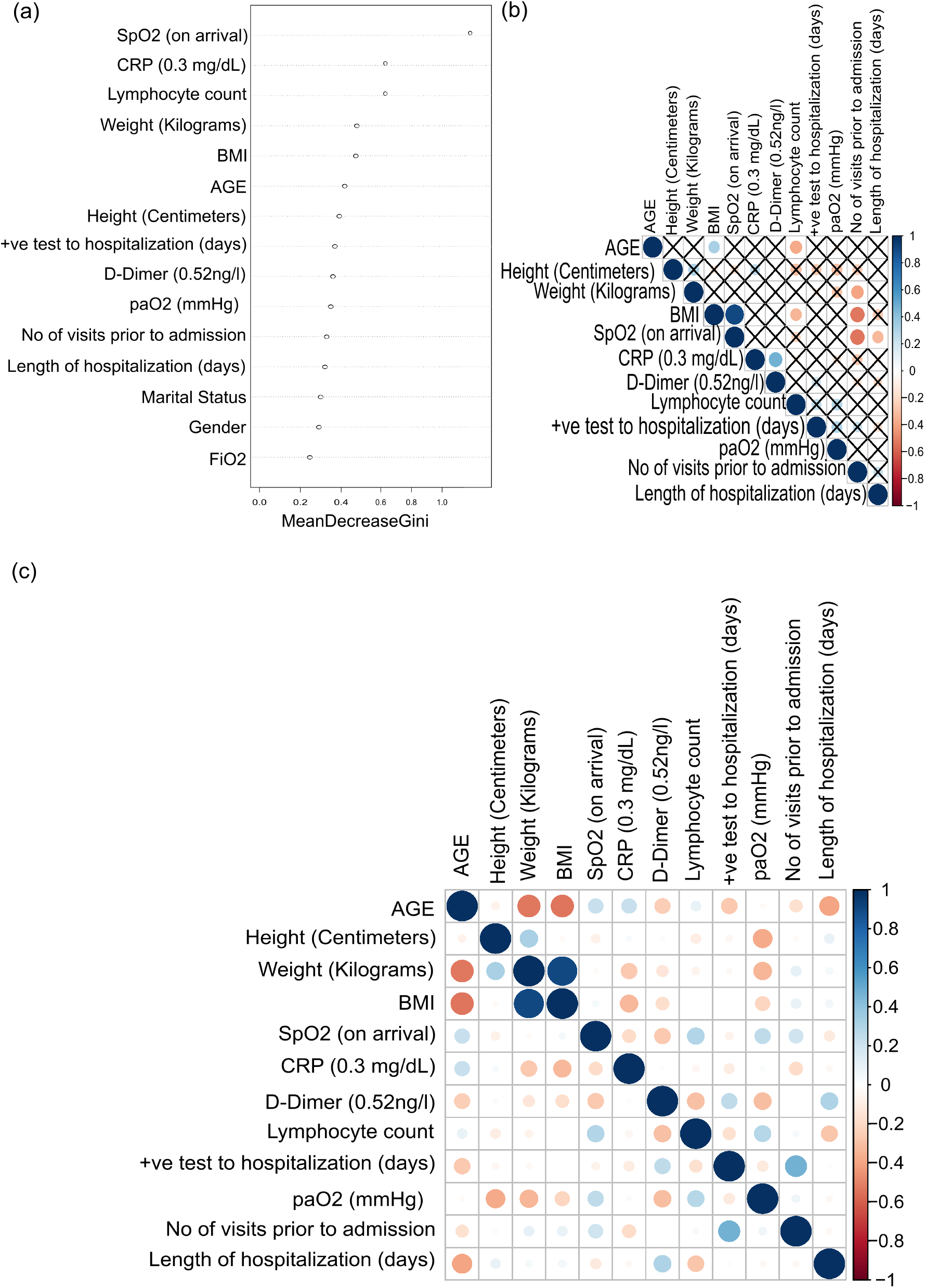
Distribution of patients based on age, height, weight, and BMI.

**Figure S2:**
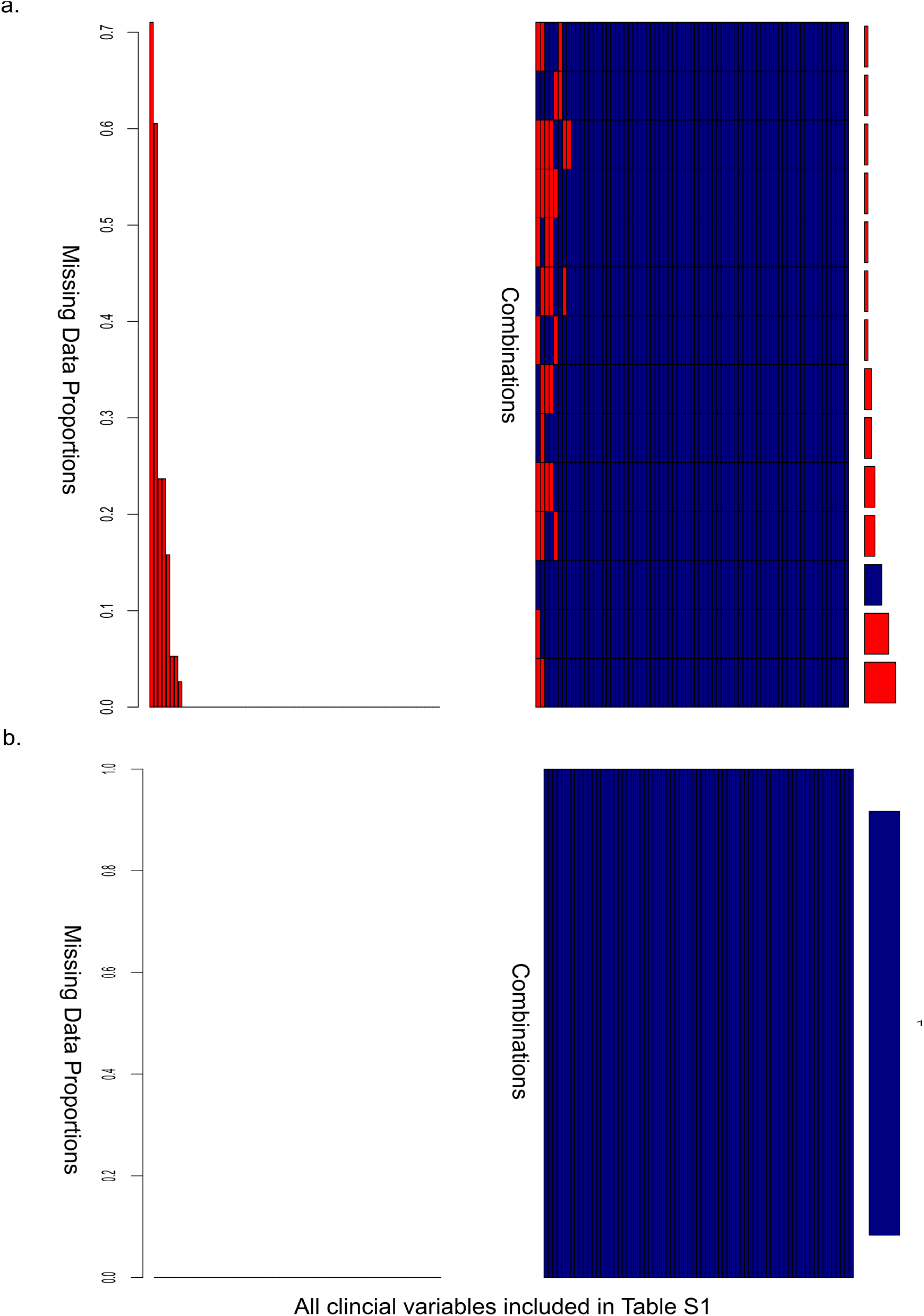
Missing values in clinical variables and imputation of missing values.

**Figure S3:**
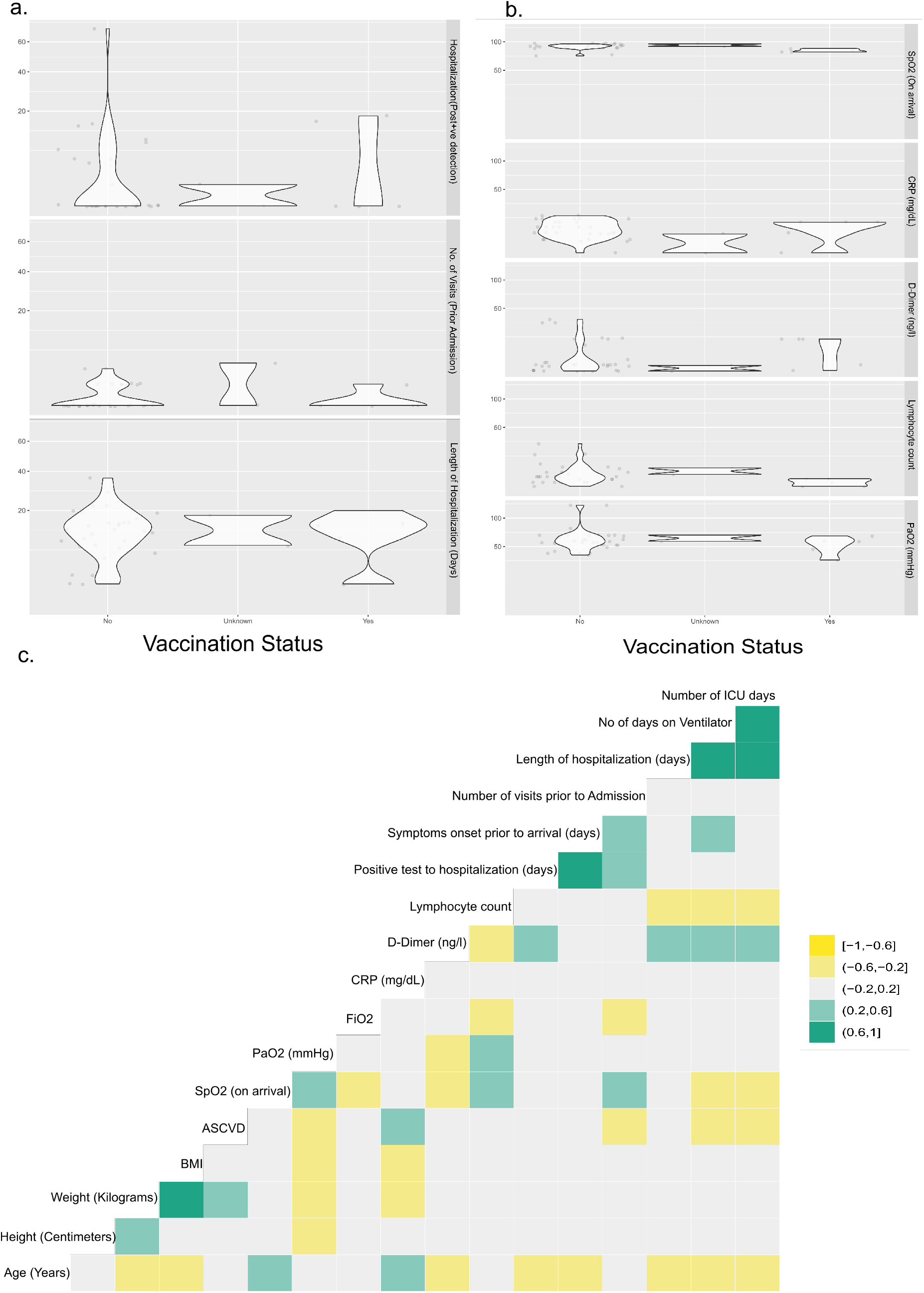
(a)Distribution of patients for vaccination status for Length of stay, patient’s RT PCR position day and hospitalization, and the total number of visits prior to hospitalization (b)Distribution of patients for vaccination status for inflammatory markers such as D-Dimer, CRP, lymphocyte and SpO2, and paO2. (c) The Pearson’s correlation of all the clinical variables in the study, the significance can be determined based on P-value <0.05.

**Figure S4:**
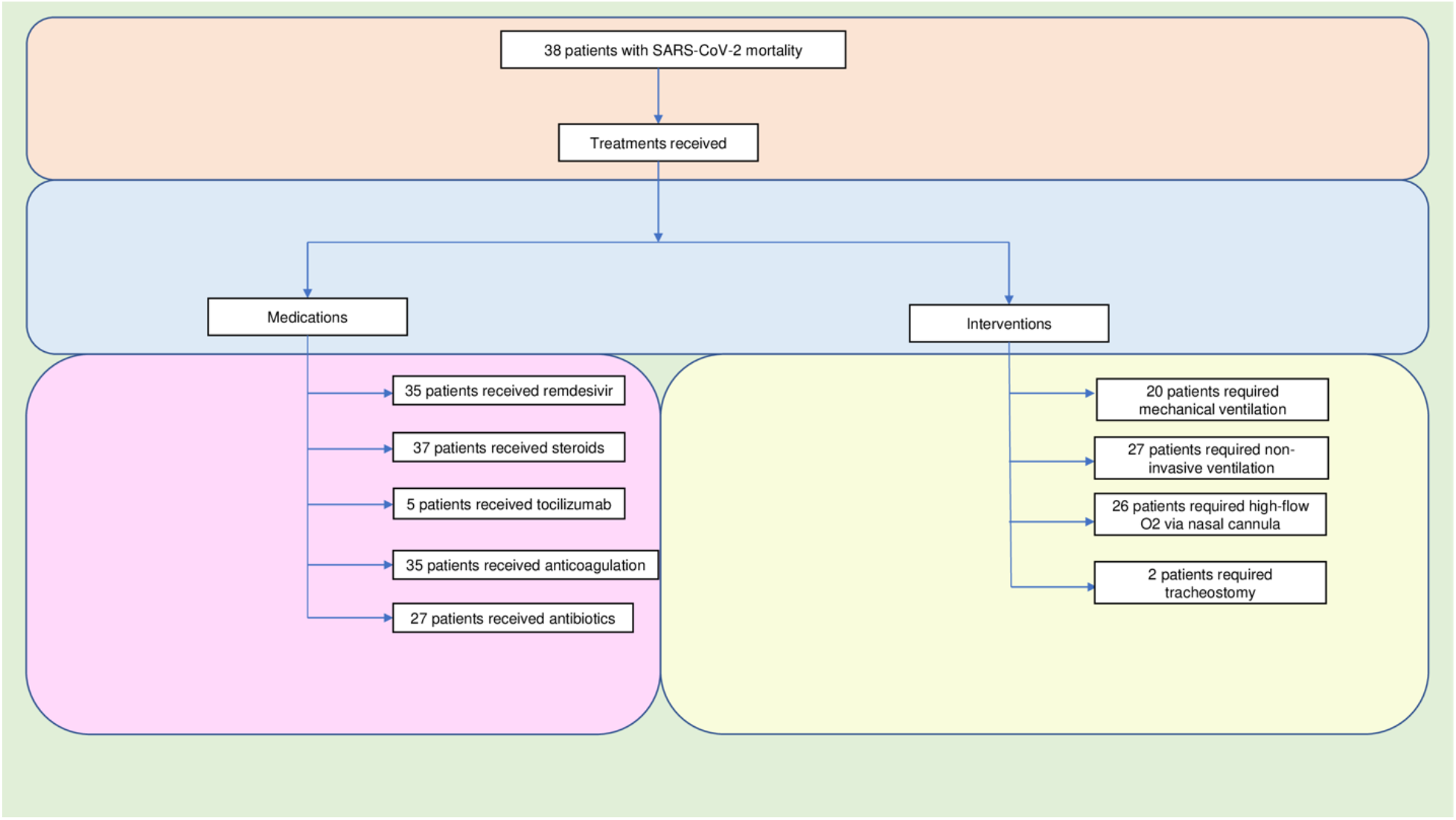
Treatment and interventions for all the patients in the cohort distributed based on medications and interventions.

**Figure S5:** The Charlson Comorbidity Index (CCI) distribution for each comorbidity for all the patients.

**Figure S6:** Each patient’s Charlson Comorbidity Index (CCI) distribution.

**Table S1:** Logistic regression to classify the vaccination status.

## Data Sharing

The data can be retrieved using the guidelines set forth by Mercy medical center.

## Contributions

NJ designed the scientific questions, formed the IRB, implemented the clinical conceptualization, identified vital datasets, and contributed to manuscript preparation. KP conceptualized, organized, and contributed to manuscript preparation and analyses results. RS, GK, and AA helped with data collection and manuscript review. Finally, AJ wrote the manuscript and helped analyze the data.

## Acknowledgments

The authors are profoundly grateful for dedicating this research to all the patients and their families unfailingly; their generosity and kindness allowed the existence of this study. In addition, the authors would like to thank Jenny Kaylor (Quality Analyst at MMC) for their help with data collection from CHI Mercy Health, Roseburg. Lastly, the authors are eternally thankful to all the healthcare and support staff involved in patient care at CHI Mercy Health, Roseburg, Oregon.

