## Supplementary material for "Meta-analysis of COVID-19 patients to understand the key predictors of mortality in the non-vaccinated groups in remote settings": Table 1

| **Characteristic** | **OR^1^** | **95% CI^1^** | **p-value** |
| --- | --- | --- | --- |
| BMI | 0.72 | 0.14, 1.47 | 0.6 |
| SpO2 (on arrival) | 0.88 | 0.62, 1.20 | 0.4 |
| CRP (mg/dL) | 0.81 | 0.55, 1.05 | 0.2 |
| D-Dimer (ng/l) | 0.93 | 0.75, 1.08 | 0.4 |
| Lymphocyte count | 0.79 | 0.34, 1.16 | 0.5 |
| Positive test to hospitalization (days) | 0.99 | 0.83, 1.10 | 0.8 |
| PaO2 (mmHg) | 0.97 | 0.84, 1.07 | 0.6 |
| Length of hospitalization (days) | 0.93 | 0.73, 1.17 | 0.5 |
| FiO2 | 1.01 | 0.95, 1.08 | 0.8 |
| ^1^OR = Odds Ratio, CI = Confidence Interval | | | |
