## Supplementary material for "Meta-analysis of COVID-19 patients to understand the key predictors of mortality in the non-vaccinated groups in remote settings": Table 2

| **Characteristic** | **N = 38^1^** |
| --- | --- |
| BMI | 31 (27, 36) |
| Vaccination status |  |
| No | 31 (82%) |
| Unknown | 2 (5.3%) |
| Yes | 5 (13%) |
| SpO2 (on arrival) | 92 (88, 94) |
| CRP (mg/dl) | 8.4 (6.1, 13.5) |
| D Dimer (ng/l) | 2 (0, 14) |
| Lymphocyte count | 4 (2, 6) |
| Positive test to hospitalization (days) | 0.0 (0.0, 4.8) |
| PaO2 (mmHg) | 58 (52, 67) |
| No of visits prior to admission |  |
| 0 | 26 (68%) |
| 1 | 9 (24%) |
| 3 | 2 (5.3%) |
| 4 | 1 (2.6%) |
| Length of hospitalization (Days) | 13 (8, 16) |
| FiO2 | 92 (41, 100) |
| ^1^Median (IQR); n (%) | |
