## Supplementary material for "Meta-analysis of COVID-19 patients to understand the key predictors of mortality in the non-vaccinated groups in remote settings": Table S1

| **Variable** | **N** | **No, N = 31^1^** | **Unknown, N = 2^1^** | **Yes, N = 5^1^** | **p-value^2^** |
| --- | --- | --- | --- | --- | --- |
| BMI | 38 | 31 (28, 35) | 36 (32, 41) | 26 (21, 35) | 0.6 |
| SpO2 (on arrival) | 38 | 92 (89, 94) | 93 (92, 94) | 82 (80, 85) | 0.006 |
| CRP (mg/dL) | 38 | 10.1 (6.1, 13.5) | 3.2 (1.7, 4.6) | 13.5 (6.7, 13.5) | 0.2 |
| D-Dimer (ng/l) | 38 | 2 (0, 7) | 1 (1, 1) | 15 (2, 15) | 0.3 |
| Lymphocyte count | 38 | 4 (2, 7) | 6 (6, 7) | 2 (1, 3) | 0.032 |
| Positive test to hospitalization (days) | 38 | 0.0 (0.0, 4.5) | 0.5 (0.2, 0.8) | 0.0 (0.0, 16.0) | 0.8 |
| PaO2 (mmHg) | 38 | 57 (52, 68) | 63 (60, 65) | 57 (46, 58) | 0.5 |
| No. of visits prior to admission | 38 |  |  |  | 0.2 |
| 0 |  | 21 (68%) | 1 (50%) | 4 (80%) |  |
| 1 |  | 8 (26%) | 0 (0%) | 1 (20%) |  |
| 3 |  | 2 (6.5%) | 0 (0%) | 0 (0%) |  |
| 4 |  | 0 (0%) | 1 (50%) | 0 (0%) |  |
| Length of hospitalization (days) | 38 | 13 (8, 16) | 13 (10, 16) | 14 (10, 15) | >0.9 |
| FiO2 | 38 | 89 (40, 100) | 54 (49, 60) | 100 (100, 100) | 0.2 |
| ^1^Median (IQR); n (%) | | | | | |
| ^2^Kruskal-Wallis rank sum test; Fisher's exact test | | | | | |
